# Asking Conversational Artificial Intelligence about Health and Subsequent Actions among Adults in Japan: A Cross-sectional Web Survey

**DOI:** 10.64898/2026.11.20.26368620

**Authors:** Mamoru Ichikawa

## Abstract

**Introduction:** People increasingly ask conversational artificial intelligence (AI) about health; however, how this relates to care seeking in Japan, where universal health coverage renders care accessible, remains unknown. This study described how many adults ask AI about health, why and when, what they do afterward, and how much they trust AI compared with other sources.

**Methods:** This two-stage cross-sectional survey employed a commercial web panel (October–November 2026). A screening survey of adults aged ≥20 years (10,000 responses; 8,209 analyzed) estimated prevalence. A main survey of users (1,000 responses; 735 analyzed) asked about purposes, the most recent use and subsequent actions, and trust in seven information sources. Responses were weighted by sex and age.

**Results:** In the past 6 months, 40.3% of adults (95% confidence interval, 39.3–41.4) asked conversational AI about health. The most recent use occurred before a medical visit for 39.0%, after a visit for 23.4%, and when no visit was being considered for 37.6%. Subsequently, 35.4% visited a medical facility; 12.8% (10.3–15.3) decided not to visit or postponed a visit (5.1% of all adults). Forgoing or delaying was more frequent among users with high trust in AI (trusting it completely or a lot; 22.5% vs. 10.9% among others). Among pre-visit users, it was less frequent with high trust in health professionals (16.7% vs. 30.9%) and did not clearly differ by trust in AI. Of users, 61.4% had high trust in health professionals and 14.3% in AI.

**Conclusions:** In Japan, approximately 4 in 10 adults asked AI about health, and 1 in 8 users forwent or delayed a visit. This was more common with high trust in AI; among pre-visit users, it was associated with low trust in health professionals. How trust in AI and health professionals relates to care seeking warrants further study.

## Introduction

Currently, people ask conversational artificial intelligence (AI) services, including ChatGPT and Gemini, about health. In the United States, 32% of adults reported using AI for information or advice about physical or mental health in the past year^1^. In Australia, 9.9% of adults surveyed in June 2024 used ChatGPT for health information in the preceding 6 months^2^. Usage records from Microsoft Copilot indicate that people also ask about family members they care for and about how to use health services^3^.

An individual who asks AI about a symptom must then decide whether to visit a clinician, wait, or manage the condition at home. How AI affects this decision bears on how health services are used. In the United States, 14% of past-30-day users said they had not seen a provider because of information from AI^4^. In the United Kingdom, 21% of individuals who had used AI for health advice had at some point decided against seeking professional advice owing to AI use^5^.

This effect may depend on trust. In four European countries, trust in AI- generated health information was moderate, and higher trust was associated with a stronger intention to use AI^6^; however, how trust relates to care seeking was not investigated. Evidence regarding how trust in AI compares with trust in health professionals, news media, search engines, and social media is limited.

These findings come from Western countries and Australia. Japan has had universal health insurance since 1961, and patients can visit almost any medical facility; in 2007, people visited physicians 13.4 times annually, more than three times as often as in the United States^7^. In the United States, more than 4 in 10 users cited cost or lack of access as reasons for using AI^1^. Whether AI use and subsequent decisions vary where care is easier to reach remains unknown. In Japan, 58.9% of individuals aged 20–69 years have used generative AI^8^; however, data on health use are limited to company surveys of specific situations, including cold symptoms^9^ and mental health^10^. To the author’s knowledge, no academic survey has estimated how many adults in Japan ask AI about health in general or described the circumstances of use and subsequent actions as proportions of the adult population.

A two-stage web survey was conducted to address four questions:

1. How many adults in Japan ask conversational AI about health, and who are they?
2. For what purposes, and at what stage of seeking care, do they use it?
3. What do they do afterward, how often do they forgo or delay a medical visit, and what is this associated with?
4. How strongly do users trust AI compared with health professionals, news media, and online sources?

## Materials and Methods

### Design and participants

A two-stage cross-sectional web survey of adults in Japan was conducted using a commercial web panel (Freeasy; iBRIDGE Corporation, Tokyo, Japan). The screening survey (October 28–29, 2026) was sent to registered panel members aged ≥20 years, with quotas set so that sex and age group matched the national population^11^, and closed at 10,000 responses. The main survey (November 9, 2026) was sent to all 3,213 screening respondents who met the quality criteria below and reported asking conversational AI about health in the past 6 months. It had no quotas and closed at 1,000 responses (31% of those invited).

Participants read an explanation of the study on the first screen of the screening survey, including that some respondents would be invited to a follow-up survey, and provided consent electronically before answering. Participants received panel points worth 5 yen (screening) and 13 yen (main survey). This study was conducted in accordance with the Declaration of Helsinki and approved by the Musashi University Research Ethics Committee (number, 2026-14; October 19, 2026). Changes made following approval and before the main survey (its sample size and the added items on trust in seven sources) were reported to the committee and accepted without re-review.

### Measures

Conversational AI was defined as “AI that you ask in sentences and that answers in sentences.” Only reading an AI summary shown in search results did not count as use; asking follow-up questions from such a summary did.

The screening survey (five items) asked which generative AI services respondents had ever used and how often they had used conversational AI for health-related questions in the past 6 months. Those reporting AI use were classified as users and were asked what they had asked about and whose health it concerned. All respondents answered a single-item health literacy question on confidence in filling out medical forms^12^.

The main survey (13 items) initially asked about the past 6 months. Purposes of use (nine items, multiple choice) and trust in what conversational AI says (5-point scale from “completely” to “not at all”) were adapted from an Australian national survey^2^, replacing “ChatGPT” with “conversational AI.” Following that survey, four purposes were classified as higher risk: learning what symptoms mean, finding out what to do about a health problem, deciding whether to see a doctor, and interpreting test results. Whether respondents consulted a health professional after using AI for physical health, and seven reasons for using AI (each rated a major reason, a minor reason, or not a reason), were adapted from a United States survey^1^. A professor of health communication who was not otherwise involved in this study checked the Japanese wording of all adapted items against the original. Respondents subsequently rated trust in seven sources of health information on the same 5-point scale: conversational AI, health professionals, television, newspapers, online news, websites found by internet search, and posts on social media or video sites.

The survey then asked about the most recent occasion of use: the service, whose health, the topic, and timing relative to a medical visit (before a visit, after a visit, or not considering a visit). Respondents selected everything they did afterward from nine actions or “did nothing in particular.” Selecting “decided not to visit” or “postponed a visit” was defined as forgoing or delaying a visit. A final item asked respondents to write, as far as they remembered, what they had typed to the AI. The complete questionnaires are presented in Supplementary Materials 1 (English translation) and 2 (Japanese original).

### Data quality and exclusions

Screening analysis excluded respondents whose mean response time was <3 s per item, a threshold that removed 503 of 595 respondents who selected the last option on every initial item, and those with a clear contradiction (reporting use of a generative AI service but selecting “I do not know conversational AI”), leaving 8,209 of 10,000.

The main analysis excluded respondents who (a) failed an attention check embedded in the reasons matrix (n = 157); (b) answered “prefer not to say” to the item on purposes of use (n = 33); (c) completed the survey in <78 s, 3 s for each of 26 response items including matrix rows (n = 135); or (d) wrote in the final item that they had not used AI for health or had typed nothing (n = 12). The criteria overlapped; 735 respondents remained (Figure 1). Criterion (a) was set before data collection; (b), (c), and (d) were fixed afterward.

**Figure 1.**
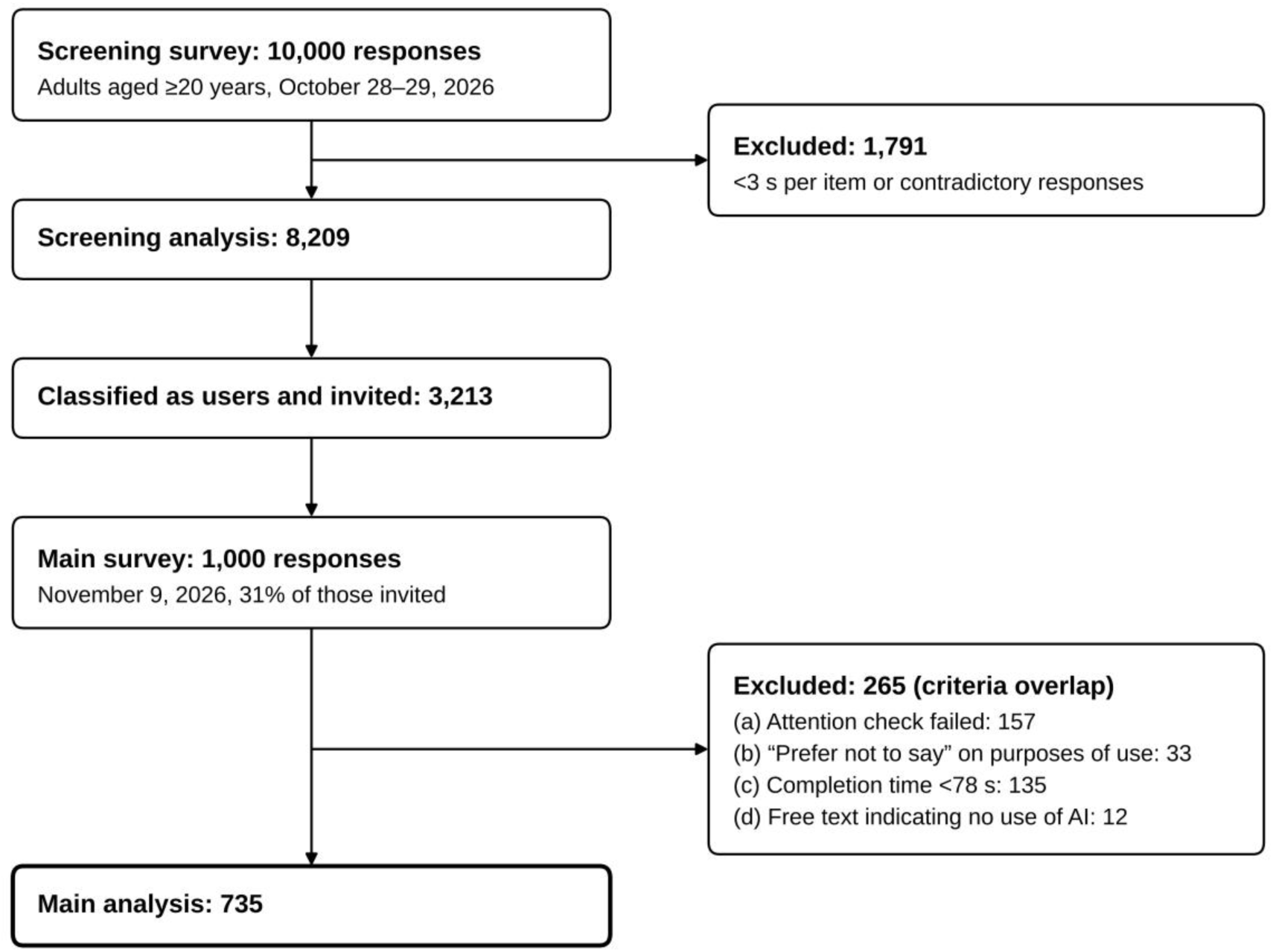
Flow of participant selection. Screening survey: 10,000 responses; 1,791 excluded (mean response time of <3 s per item or contradictory responses); 8,209 respondents analyzed, 3,213 of whom were classified as users and invited to the main survey. Main survey: 1,000 responses; 265 excluded under criteria (a) attention check failed (n = 157), (b) “prefer not to say” to the item on purposes of use (n = 33), (c) completion time of <78 s (n = 135), and (d) free text indicating no use of AI (n = 12); 735 analyzed. Numbers are unweighted. The criteria overlap; therefore, the numbers by criterion do not sum to the number excluded. AI, artificial intelligence

### Weighting

Exclusions were concentrated among younger respondents (>50% of those in their 20s, <4% of those aged ≥70 years). Screening responses were post-stratified to the sex-by-age distribution (six age groups) of the Japanese population aged ≥20 years as of October 1, 2025^13^. Main-survey responses were weighted to the sex-by-age distribution of weighted screening users. Main-analysis weights were 0.71–2.12; the effective sample size^14^ was 664.

### Statistical analysis

Weighted percentages were reported with 95% confidence intervals (CIs) based on the effective sample size. Moreover, actions were expressed as shares of all adults, by multiplying prevalence by the share of users taking the action; for this, prevalence was corrected for users identified by criterion (d) as not having used AI. CIs for these shares were obtained using the delta method. The association between health literacy and use was investigated using survey-weighted logistic regression (svyglm) adjusted for sex and age.

For the seven sources, the reported measures were the percentage trusting each source “completely” or “a lot” and the standardized within-person difference from conversational AI (Cohen’s dz, the weighted mean of within-respondent differences divided by their weighted standard deviation). Differences in percentages were tested by comparing the weighted mean of within-respondent differences with zero (svymean), with Bonferroni correction for six comparisons. “Don’t know” and “prefer not to say” were treated as missing for scale scores.

An exploratory survey-weighted logistic regression (svyglm) was fitted to describe factors associated with forgoing or delaying a visit. Predictors were sex, age group, timing relative to a visit, cost as a reason for use, any mental health-related question (in either survey), trust in conversational AI, trust in health professionals, and the number of purposes selected. As forgoing a visit presupposes considering one, the tabulations and regression were repeated among those who used AI before a visit (n = 284).

As sensitivity analyses, the main estimates and regression were repeated under six alternative sets of exclusion criteria; prevalence was recalculated under four alternative screening criteria, including time thresholds of 2 and 4 s. Analyses were performed using R 4.6.2 (R Foundation for Statistical Computing, Vienna, Austria) with the survey package. Reporting followed the STROBE statement^15^ and CHERRIES^16^.

### Generative AI use

The author designed the study and questionnaires, decided the exclusion criteria and analyses, and interpreted the results. Under the author’s direction, Claude (Anthropic; Claude Opus 5.5 and Claude Fable 5.2) assisted in writing the R code, creating figures from the results, and drafting and editing the manuscript.

## Results

### Prevalence

Of the 8,209 screening respondents analyzed, 40.3% (95% CI, 39.3–41.4) had asked conversational AI about health in the past 6 months (Table 1). As some respondents initially classified as users proved in the main survey not to have used AI (criterion [d]; weighted 1.7%), the corrected prevalence was 39.7%. Under alternative screening criteria, prevalence ranged from 36.5% (treating 320 respondents whose free-text responses described non-health uses as nonusers) to 41.1% (time threshold of 4 s per item); it was 37.2% with no exclusions.

**Table 1.** Adults who asked conversational AI about health in the past 6 months (screening survey, n = 8,209)

|  | **Unweighted n** | **Asked conversational AI about health, % (95% CI)** | **Ever used generative AI, % (95% CI)** | **Asked about health, among those who ever used generative AI, % (95% CI)** |
|---|---|---|---|---|
| All | 8,209 | 40.3 (39.3–41.4) | 53.0 (51.9–54.1) | 73.8 (72.4–75.1) |
| Males | 3,965 | 44.0 (42.4–45.6) | 57.7 (56.1–59.3) | 74.5 (72.6–76.3) |
| Females | 4,244 | 36.9 (35.5–38.4) | 48.7 (47.2–50.2) | 73.0 (71.0–75.0) |
| 20–29 years | 571 | 49.4 (45.3–53.5) | 68.9 (65.1–72.7) | 71.0 (66.6–75.5) |
| 30–39 years | 790 | 50.0 (46.5–53.5) | 64.4 (61.1–67.8) | 76.0 (72.3–79.7) |
| 40–49 years | 1,199 | 48.1 (45.3–51.0) | 61.3 (58.5–64.1) | 77.0 (74.0–80.0) |
| 50–59 years | 1,562 | 41.5 (39.1–44.0) | 54.8 (52.4–57.3) | 74.1 (71.2–77.0) |
| 60–69 years | 1,369 | 35.9 (33.4–38.5) | 47.9 (45.3–50.6) | 72.4 (69.0–75.8) |
| ≥70 years | 2,718 | 30.0 (28.2–31.7) | 39.0 (37.2–40.8) | 71.9 (69.2–74.6) |

Percentages are weighted. AI, artificial intelligence; CI, confidence interval.

Younger individuals and males more frequently asked AI about health; however, the difference arose at adoption of generative AI: among adopters, 71%–77% used it for health in every age group and both sexes. On the health literacy item, 17.9% were “not at all confident” with medical forms; their use was 25.0% (22.6–27.3), compared with 43.7% (42.5–44.9) among other respondents (adjusted odds ratio, 0.35; 95% CI, 0.31– 0.41), and use did not vary across the other four levels (43%–44%); this held under the alternative screening criteria.

Main-survey response was higher among males (35.2% vs. 26.4%) and lowest among those aged ≥70 years (20.8%). Weekly or more frequent use at screening was comparable in respondents (47.3%) and nonrespondents (43.2%).

### Purposes and stage of seeking care

Learning what symptoms mean (52.8%) was the most common purpose of use; 63.3% used AI for at least one higher-risk purpose (Table 2), comparable to the 61% reported in Australia with the same item^2^. The most recent use, most frequently using ChatGPT (40.3%) or Gemini (35.1%), occurred before a medical visit for 39.0%, after a visit for 23.4%, and when no visit was being considered for 37.6%.

**Table 2.** Characteristics of users and their use of conversational AI for health (main survey, n = 735)

| Item | Weighted % (95% CI) |
|---|---|
| **Sex** |  |
| Females (n = 297) | 47.5 (43.7–51.3) |
| Males (n = 438) | 52.5 (48.7–56.3) |
| **Age** |  |
| 20–29 years (n = 54) | 13.9 (11.3–16.6) |
| 30–39 years (n = 98) | 15.1 (12.4–17.8) |
| 40–49 years (n = 150) | 18.3 (15.3–21.2) |
| 50–59 years (n = 171) | 18.4 (15.5–21.4) |
| 60–69 years (n = 116) | 13.0 (10.5–15.6) |
| ≥70 years (n = 146) | 21.3 (18.2–24.4) |
| **Purposes of use, past 6 months (multiple choice)** |  |
| Learn what symptoms mean* | 52.8 (49.0–56.6) |
| Learn about a disease or health condition | 36.4 (32.7–40.1) |
| Learn about medicines, tests, or treatments | 29.6 (26.1–33.0) |
| Find out what to do about a health problem* | 26.8 (23.4–30.1) |
| Learn about healthy lifestyle habits | 23.6 (20.4–26.8) |
| Understand medical terms | 18.5 (15.5–21.4) |
| Make a plan to improve health | 15.0 (12.3–17.7) |
| Decide whether to see a doctor* | 11.8 (9.3–14.2) |
| Interpret test results* | 11.4 (9.0–13.8) |
| At least one higher-risk purpose (*) | 63.3 (59.6–66.9) |
| **Most recent use: service** |  |
| ChatGPT | 40.3 (36.5–44.0) |
| Gemini | 35.1 (31.4–38.7) |
| AI Mode in Google Search | 17.7 (14.8–20.6) |
| Copilot | 4.4 (2.8–5.9) |
| Claude | 1.1 (0.3–1.9) |
| Other | 1.5 (0.6–2.4) |
| **Most recent use: whose health** |  |
| Own | 86.2 (83.6–88.8) |
| Child | 3.5 (2.1–4.9) |
| Parent | 4.4 (2.9–6.0) |
| Other family member | 4.7 (3.1–6.3) |
| Other | 1.2 (0.4–2.0) |
| **Most recent use: topic** |  |
| Acute symptom | 36.9 (33.2–40.6) |
| Chronic condition | 18.9 (15.9–21.9) |
| Test results | 16.2 (13.4–19.0) |
| Medicines | 13.3 (10.7–15.8) |
| Mental health | 12.2 (9.8–14.7) |
| Other | 2.5 (1.3–3.6) |
| **Most recent use: timing relative to a medical visit** |  |
| Before a visit | 39.0 (35.3–42.7) |
| After a visit | 23.4 (20.1–26.6) |
| Not considering a visit | 37.6 (33.9–41.3) |

n is unweighted. CIs are based on the effective sample size. \*Purposes classified as higher risk by Ayre et al.<sup>2</sup>. Multiple-choice items do not sum to 100%. AI, artificial intelligence; CI, confidence interval.

Wanting information or support right away (71.9%) and wanting to examine the matter before deciding whether to seek care (64.5%) represented the leading reasons for use. The seven reasons followed almost the same order as in the United States (Spearman’s rho = 0.96)^1^. The proportions citing being unable to afford care (23.9% vs. 46%) and having no regular provider or being unable to get an appointment (26.6% vs. 44%) were lower than those in the United States.

### Actions after use

Following the most recent use, 35.4% of users visited a medical facility, and 41.4% did nothing in particular (Table 3). Among those who had used AI for physical health in the past 6 months (n = 620), 37.8% (33.8–41.8) subsequently consulted a health professional, compared with 58% in the United States^1^. Of those who visited following the most recent use (n = 259), 10.9% (6.9–14.9) told a physician or pharmacist what they had learned using AI.

**Table 3.** Actions after the most recent use of conversational AI for health (main survey, n = 735)

| **Item** | **Weighted % (95% CI)** |
|---|---|
| **All users (multiple choice)** |  |
| Visited a medical facility | 35.4 (31.7–39.0) |
| Decided not to visit | 6.2 (4.4–8.0) |
| Postponed a visit | 8.6 (6.5–10.7) |
| Forwent or delayed a visit (either of the two above) | 12.8 (10.3–15.3) |
| Bought or used an over-the-counter medicine | 11.8 (9.3–14.2) |
| Changed or stopped how a prescription medicine was taken | 2.9 (1.6–4.2) |
| Told a physician or pharmacist what was learned using AI | 5.6 (3.9–7.4) |
| Called a telephone health advice line (#7119 or #8000) | 2.2 (1.0–3.3) |
| Called an ambulance or visited an emergency department | 1.1 (0.3–1.8) |
| Did nothing in particular | 41.4 (37.6–45.1) |
| **Used AI before a visit (n = 284)** |  |
| Visited a medical facility | 64.1 (58.2–69.9) |
| Forwent or delayed a visit | 21.3 (16.3–26.4) |
| Did nothing in particular | 12.3 (8.2–16.3) |
| **Used AI after a visit (n = 169)** |  |
| Visited a medical facility | 41.4 (33.5–49.2) |
| Forwent or delayed a visit | 8.0 (3.7–12.3) |
| Did nothing in particular | 35.8 (28.2–43.4) |
| **Not considering a visit (n = 282)** |  |
| Visited a medical facility | 1.9 (0.2–3.6) |
| Forwent or delayed a visit | 7.0 (3.8–10.1) |
| Did nothing in particular | 75.0 (69.7–80.3) |

n is unweighted. CIs are based on the effective sample size. AI, artificial intelligence; CI, confidence interval.

Overall, 12.8% (10.3–15.3) of users decided not to visit or postponed a visit, corresponding to 5.1% (4.1–6.1) of all adults. Users who used AI before a visit accounted for 65.0% of those who forwent or delayed.

Forgoing or delaying was reported by 22.5% (14.4–30.6) of users who trusted what conversational AI says completely or a lot on the single trust item and 10.9% (8.3– 13.4) of other users. However, high-trust users more frequently used AI before a visit (52.7% vs. 36.3%). Among the 284 who used AI before a visit, no clear difference was observed by trust in AI (26.8% vs. 19.7%), whereas a difference was noted by trust in health professionals (16.7% among those trusting health professionals completely or a lot vs. 30.9% among others).

In the exploratory regression (Table 4), forgoing or delaying was associated with pre-visit use, citing cost as a reason, and lower trust in health professionals. Moreover, it was more frequent among users who had asked about mental health (27.2% of users; forgoing or delaying, 19.9% vs. 10.2%). Trust in conversational AI, sex, age, and number of purposes demonstrated no clear association. In the regression restricted to pre-visit users (n = 280), the association with trust in health professionals remained (odds ratio per level, 0.60; 0.42–0.86), whereas trust in AI showed no association (1.04; 0.64–1.71).

**Table 4.**
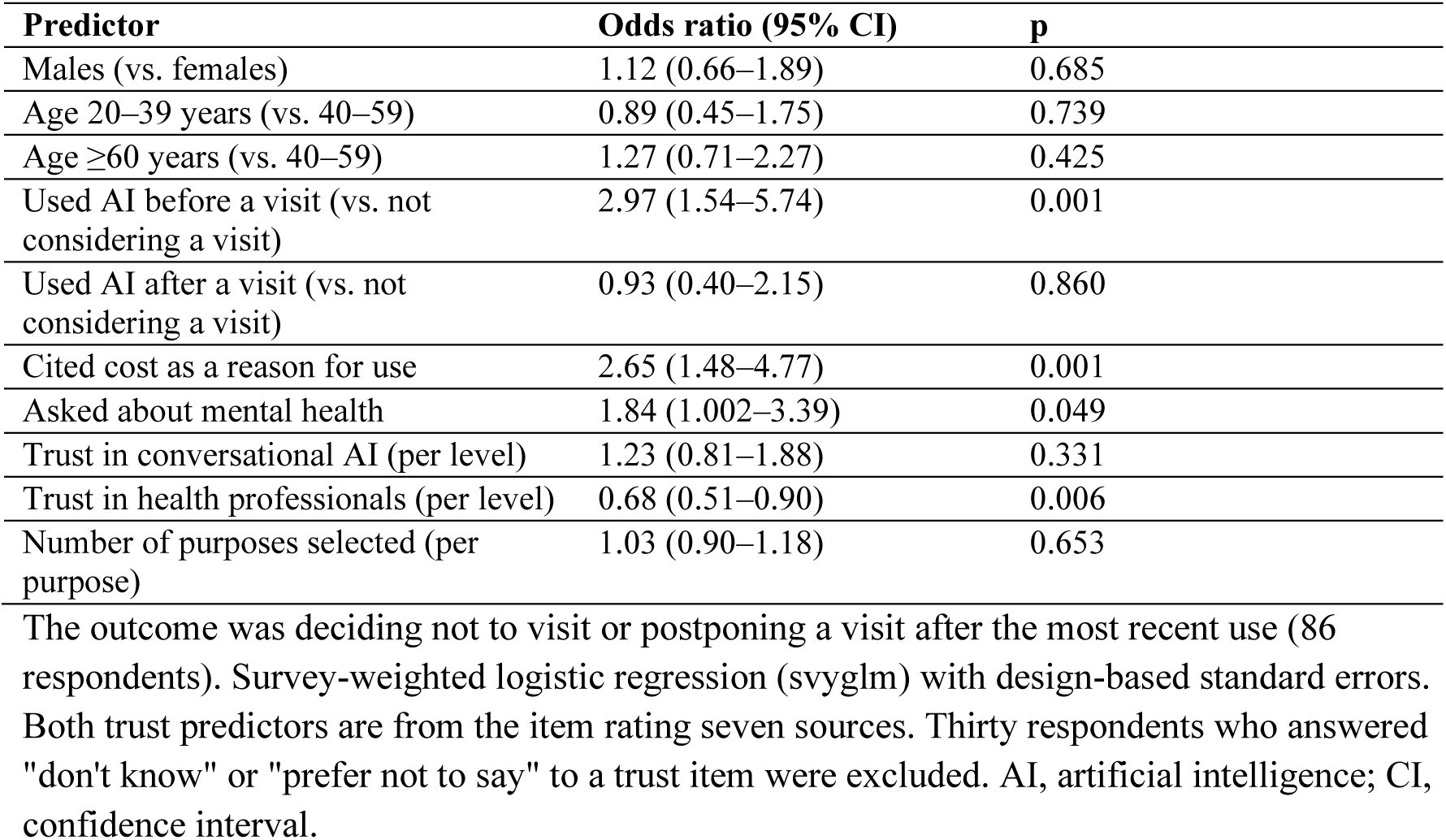
Factors associated with forgoing or delaying a medical visit after the most recent use (exploratory logistic regression, n = 705)

### Trust in AI and other sources

On the single trust item, 16.7% of users trusted what conversational AI says “completely” or “a lot,” 47.8% “somewhat,” and 31.8% “a little” or “not at all” (mean, 2.83 on the 5-point scale; 3.1 in Australia^2^).

Across the seven sources (Figure 2), 61.4% of users trusted health professionals completely or a lot, more than any other source. Conversational AI (14.3%) did not differ from television (16.1%); was lower than newspapers (20.9%); and was higher than online news, websites found by search, and social media (all <10%). Regarding mean scores, conversational AI (2.77) did not differ from newspapers (2.81; dz = −0.04) or television (2.68; dz = 0.09), was lower than health professionals (3.67; dz = −0.96), and was higher than the three online sources (dz = 0.43–0.84). Of the 705 respondents who rated both AI and health professionals, 4.8% trusted AI more than health professionals.

**Figure 2.**
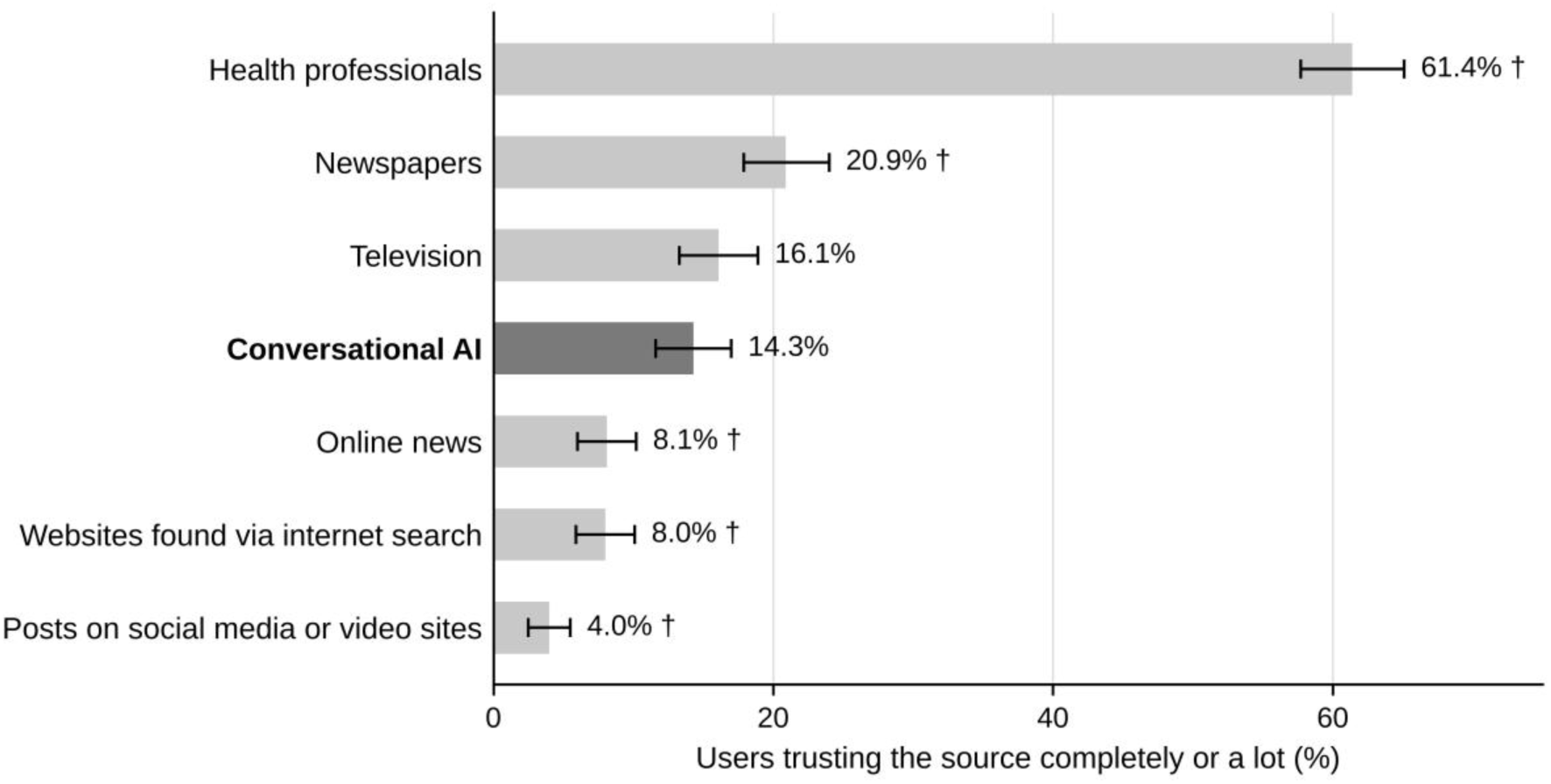
Percentage of users who trust each source of health and medical information “completely” or “a lot” (n = 735) Health professionals, 61.4% (57.7–65.1)†; newspapers, 20.9% (17.9–24.0)†; television, 16.1% (13.3–18.9); conversational AI, 14.3% (11.6–17.0); online news, 8.1% (6.0– 10.2)†; websites found via internet search, 8.0% (5.9–10.1)†; and posts on social media or video sites, 4.0% (2.5–5.5)†. Weighted percentages with 95% confidence intervals based on the effective sample size. The denominator includes “don’t know” and “prefer not to say.” †Significantly different from conversational AI (weighted within-respondent test; Bonferroni-corrected p < 0.05 for six comparisons). AI, artificial intelligence

### Sensitivity analyses

Across descriptive estimates, the largest change was 3.4 percentage points when the time threshold was varied and 9.6 points with no exclusions, both for wanting information or support right away (Table 5). The associations of forgoing or delaying with pre-visit use and trust in health professionals held under the main criteria and all six alternatives. The association with cost was no longer clear at 4 s per item, whereas that with mental health-related questions held under four of the seven sets. Trust in AI was associated with the outcome only when no exclusions were applied.

**Table 5.** Sensitivity analyses: main estimates and regression results under alternative exclusion criteria.

|  | **Main analysis** | **No exclusions** | **Criteria (a) and (b) only** | **Without criterion (d)** | **Time threshold 2 s per item** | **Time threshold 4 s per item** | **Also excluding identical ratings of all 7 sources** |
|---|---|---|---|---|---|---|---|
| n | 735 | 1,000 | 824 | 747 | 793 | 636 | 686 |
| Visited a medical facility, % | 35.4 | 31.9 | 34.0 | 34.7 | 34.8 | 36.1 | 35.7 |
| Forwent or delayed a visit, % | 12.8 | 13.5 | 13.4 | 12.5 | 13.3 | 12.4 | 12.0 |
| Used AI before a visit, % | 39.0 | 36.3 | 38.6 | 38.4 | 39.3 | 37.6 | 39.3 |
| At least one higher-risk purpose, % | 63.3 | 55.4 | 61.5 | 62.4 | 62.6 | 64.6 | 62.9 |
| Trust conversational AI completely or a lot (seven-source item), % | 14.3 | 15.0 | 15.2 | 14.1 | 14.8 | 15.0 | 14.0 |
| Trust health professionals completely or a lot, % | 61.4 | 56.1 | 60.3 | 61.3 | 60.7 | 63.0 | 64.6 |
| Wanted information or support right away (reason for use), % | 71.9 | 62.3 | 70.6 | 70.8 | 71.2 | 75.3 | 72.3 |
| n in regression | 705 | 940 | 791 | 716 | 762 | 613 | 662 |
| OR: used AI before a visit | 2.97 (1.54–5.74) | 2.63 (1.55–4.47) | 3.34 (1.84–6.06) | 3.08 (1.60–5.94) | 3.03 (1.66–5.55) | 3.79 (1.77–8.10) | 2.81 (1.41–5.63) |
| OR: cited cost as a reason | 2.65 (1.48–4.77) | 2.69 (1.66–4.36) | 2.84 (1.68–4.80) | 2.66 (1.48–4.78) | 3.00 (1.76–5.11) | 1.75 (0.87–3.50) | 2.69 (1.43–5.05) |
| OR: asked about mental health | 1.84 (1.002–3.39) | 1.15 (0.70–1.88) | 1.52 (0.88–2.62) | 1.84 (1.00–3.37) | 1.69 (0.96–2.97) | 2.06 (1.03–4.10) | 2.15 (1.15–4.02) |
| OR: trust in conversational AI | 1.23 (0.81–1.88) | 1.60 (1.17–2.19) | 1.40 (0.97–2.00) | 1.24 (0.81–1.89) | 1.42 (0.96–2.10) | 1.20 (0.73–1.96) | 1.20 (0.76–1.89) |
| OR: trust in health professionals | 0.68 (0.51–0.90) | 0.63 (0.50–0.79) | 0.65 (0.50–0.83) | 0.68 (0.51–0.89) | 0.66 (0.51–0.85) | 0.67 (0.49–0.92) | 0.72 (0.54–0.97) |

Percentages are weighted, with weights recalculated under each set of criteria. Odds ratios (95% confidence intervals) are from the model in Table 4. Exclusion criteria (a) to (d) are defined in the Methods. AI, artificial intelligence; OR, odds ratio.

## Discussion

Approximately 4 in 10 adults in this survey had asked conversational AI about health in the past 6 months. Approximately 1 in 8 users decided not to visit or postponed a visit following their most recent AI use. Users trusted AI far less than health professionals and about as much as television and newspapers.

### Prevalence

The 39.7%–40.3% prevalence is higher than the 32% reported for the past year in the United States^1^ and approximately four times the 9.9% reported for ChatGPT in Australia in 2024^2^. These surveys vary in timing, services covered, and sampling, and members of an opt-in web panel are likely more active online than the general public.

Younger individuals and males more frequently used AI for health because they more frequently used generative AI at all; among adopters, approximately three-quarters used it for health regardless of age or sex. By contrast, individuals not at all confident with medical forms used it less. In Australia, individuals with limited health literacy were more likely to use ChatGPT for health-related queries^2^, the opposite direction.

### Purposes and reasons

Approximately two-thirds of users asked higher-risk questions, almost the same share as in Australia. The order of reasons was similar to that in the United States; however, cost and access were cited approximately half to three fifths as often. This is expected under universal insurance with free choice of provider; however, approximately a quarter of users still provided these reasons.

### Actions after use

Approximately two-thirds of pre-visit users went on to visit a medical facility, whereas most who were not considering a visit did nothing further. However, the proportion of users who consulted a health professional after using AI was lower in Japan than in the United States. The reference periods vary (Japan, 6 months; United States, 1 year); however, easy access to care did not translate into more professional consultation following AI use.

Forgoing or delaying a visit was not rare: 12.8% of users, or approximately 5% of all adults, based on one most recent occasion. Surveys with different questions report comparable values: 14% in the United States^4^ and 21% in the United Kingdom^5^. Whether these decisions were appropriate was not assessed.

Forgoing or delaying was approximately twice as frequent among users with high trust in AI. However, these users more frequently used AI before a visit; among pre-visit users, the clear difference was by trust in health professionals, not trust in AI. Before using AI, users who forwent or delayed may already have been hesitant to seek care owing to low trust in professionals or cost. The data are cross-sectional, and the predictors were selected after data collection; therefore, causality cannot be shown. Only 60 pre-visit users had high trust in AI; therefore, a slight difference may have been missed.

### Trust in AI and other sources

Users did not treat AI as an authority: 1 in 6 trusted it completely or a lot, and fewer than 1 in 20 trusted it more than health professionals. Regarding mean scores, AI was comparable to television and newspapers and higher than online news, search results, and social media; however, more users expressed high trust in newspapers than in AI. This comparison applies to users alone; nonusers, who were not asked, may trust AI less.

### Implications

First, clinicians are likely unaware of most AI use by their patients: only approximately 1 in 10 users who visited told a physician or pharmacist what they had learned using AI. Asking patients whether they consulted AI, and what it said, could bring information that is already influencing their decisions into the consultation.

Second, forgoing or delaying among pre-visit users was associated with low trust in health professionals; it was more common among those who cited cost. Efforts to reduce harm from AI use should address not only the accuracy of AI but also access to care and trust in health professionals.

Third, television and radio are the most frequently cited sources of medical information in Japan (37.2% in a 2024 Tokyo survey; newspapers, 16.3%)^17^. Among users, AI already receives about the same trust as television and newspapers, and it answers each person’s question directly. Health journalism can no longer regard disseminating new findings to the public as its sole function, and what role it should play requires reconsideration.

### Limitations

Panel members were not randomly selected from the adult population; therefore, the results cannot be directly generalized to all adults in Japan. Weighting corrected only for sex and age, and individuals aged ≥85 years were almost absent; therefore, prevalence is probably overestimated. Among screening respondents, >50% in their 20s were excluded owing to very short response times; estimates for this group rely on few respondents, who may differ from those excluded. The boundary of “use” was not uniform: some respondents appeared to use AI through search engines without regarding it as generative AI, and approximately 1 in 10 screening users described non-health-related uses or no use in free text.

All measures were self-reported. Actions were asked about one most recent occasion, which may be imperfectly recalled and underestimates the share who ever forwent or delayed a visit during the period. The main survey ended when responses reached the target; therefore, early responders may be overrepresented. Some exclusion criteria and regression predictors were decided after data collection; therefore, results under alternative criteria were reported.

## Conclusion

In Japan, where care is comparatively accessible, approximately 4 in 10 adults asked AI about health, and most pre-visit users went on to visit a medical facility. Approximately 1 in 8 users forwent or delayed a visit. Forgoing or delaying was more frequent among high-trust users; however, among pre-visit users, it was associated with low trust in health professionals. How users’ trust in AI and in health professionals relates to care-seeking decisions following AI use warrants further study.

## Supporting information

Supplementary Materials 1 and 2: Questionnaires (English translation and Japanese original)

## Author contributions

Mamoru Ichikawa conceived and designed the study, collected and analyzed the data, interpreted the results, and wrote the manuscript.

## IRB approval code and institution

No. 2026-14, Research Ethics Committee of Musashi University (approved October 19, 2026)

## Acknowledgments

The author thanks Professor Hirono Ishikawa (Teikyo University Graduate School of Public Health) for checking the Japanese wording of the items adapted from prior surveys.

## Use of generative AI

Claude (Anthropic; Claude Opus 5.5 and Claude Fable 5.2) was used to assist in writing the R code for the analyses that the author ran and checked, and in drafting and editing the text, tables, and figures in English. The author directed each step, checked the outputs against the data and original sources, and takes full responsibility for the content.

## Data availability

The questionnaires are provided in Supplementary Materials 1 and 2. Individual responses are not shared, as participants were told that results would be published only in aggregate.

