## Supplementary Materials 1 and 2: Questionnaires (English translation and Japanese original) for "Asking Conversational Artificial Intelligence about Health and Subsequent Actions among Adults in Japan: A Cross-sectional Web Survey"

### Supplementary Material 1. Questionnaires (English translation)

This translation was prepared for reporting; both surveys were administered in Japanese (Supplementary Material 2). Items adapted from published surveys use the original English wording, with “ChatGPT” or “AI tool or chatbot” replaced by “conversational AI.” All items were mandatory, and options were shown in a fixed order. The consent text is not included. Notes in square brackets were not shown to respondents.

#### Screening survey (October 28-29, 2026; 5 items)

**Q1.** Which of the following have you ever used? (Select all that apply.)

- ChatGPT (OpenAI)
- Copilot (Microsoft)
- Gemini (Google)
- Other generative AI (please write the name of the service)
- I have never used any of these

[The Japanese screen also showed each service name in katakana.]

Text shown before Q2: The following questions are about “conversational AI.” Conversational AI means AI, such as ChatGPT, Gemini, Copilot, and Claude, that you ask in sentences and that answers in sentences. Only reading an AI summary shown in search results does not count. Asking follow-up questions from an AI summary (such as AI Mode in Google Search) does count.

**Q2.** In the past 6 months, how often have you used conversational AI to get answers to questions about health? (Select one.)

1. A few times (about 1 to 5 times)
2. Once a month
3. Once a week
4. More than once a week
5. I do not know what conversational AI is
6. Not in the past 6 months, but 6 months to 1 year ago
7. Not for more than 1 year (including never)
8. Don’t know
9. Prefer not to say

[Respondents who selected 1 to 4 were classified as users.]

**Q3.** If you need to go to the doctor, clinic or hospital, how confident are you filling out medical forms by yourself? (Select one.)

- Extremely
- Quite a bit
- Somewhat
- A little bit
- Not at all

[Single-item health literacy screener (reference 12).]

**Q4.** In the past 6 months, what did you ask conversational AI about? (Select all that apply.) [Users only.]

- Sudden symptoms (fever, pain, injury, etc.)
- A chronic condition (an ongoing illness)
- Mental health problems (low mood, anxiety, trouble sleeping, etc.)
- Test results (health checkups, blood tests, imaging, etc.)
- Medicines (how to take them, side effects, interactions, etc.)
- Other (please specify)

**Q5.** In the past 6 months, whose health did you ask conversational AI about? (Select all that apply.) [Users only.]

- Myself
- My child
- My parent
- Another family member
- Other (a friend, acquaintance, colleague, etc.; please specify)

#### Main survey (November 9, 2026; 13 items)

Text shown before Q1: First, we ask about questions you asked conversational AI in the past 6 months about your own health or the health of someone close to you.

**Q1.** In the past 6 months, have you used conversational AI to do any of the following? (Select all that apply.)

- Find out what my symptoms mean (or the symptoms of someone I know)
- Find out what to do about a specific health issue that I or someone I know has
- Learn about a specific health condition
- Learn about healthy lifestyles
- Help create a plan to improve my health (or the health of someone I know)
- Learn more about a medicine, test or treatment (eg, safety, side effects or interactions)
- Find out if I or someone I know should see a doctor

- Understand medical terms
- Interpret results from blood tests or imaging
- Other
- Don’t know (cannot be selected with other options)
- Prefer not to say (cannot be selected with other options)

[Adapted from Ayre et al. (reference 2).]

**Q2.** How much do you trust the answers that conversational AI gives? (Select one.)

- Completely
- A lot
- Somewhat
- A little
- Not at all
- Don’t know
- Prefer not to say

[Adapted from Ayre et al. (reference 2).]

**Q3.** For health and medical information, how much do you trust what each of the following tells you? (Select one for each item.)

Items, shown in this order:

- Answers given by conversational AI
- Explanations by physicians and other health professionals
- Television (news and health programs)
- Newspaper articles (print or online)
- Online news (such as Yahoo! News)
- Websites found by internet search
- Posts on social media or video sites (such as X and YouTube)

[Options are the same seven as in Q2. Developed for this study.]

**Q4.** After using conversational AI for information or advice related to your physical health (excluding mental health), did you follow up with a doctor or other health professional? (Select one.)

- Yes, followed up
- No, did not follow up
- I have not used it for physical health

[Adapted from KFF (reference 1).]

**Q5.** People use conversational AI for health information for different reasons. For each of the following, please indicate whether it was a major reason, a minor reason, or not a reason you used conversational AI for health information. (Select one for each item.)

Items, shown in this order:

- You don't have a regular health care provider, or could not get an appointment
- You could not afford the cost of seeing a provider
- You wanted to look up information before deciding whether to see a health care provider
- You felt more comfortable looking up things related to your health privately
- For this item, please select "a minor reason" [attention check]
- You felt the information from conversational AI was as reliable as what a health care provider would tell you
- You received medical test results before being able to discuss them with a provider
- You wanted quick or immediate information or support

[Options: a major reason, a minor reason, not a reason. Adapted from KFF (reference 1), except the fifth item.]

Text shown before Q6: The following questions are about the most recent time you asked conversational AI about health.

**Q6.** Which service did you mainly use that time? (Select one.)

- ChatGPT
- Gemini (app or website)
- Copilot
- Claude
- AI Mode in Google Search (including follow-up questions from an AI summary in search results)
- Other (free text)

**Q7.** Whose health was the question about? (Select one.)

- Myself
- My child
- My parent
- Another family member
- Other (a friend, acquaintance, etc.; free text)

**Q8.** What was the question about? (Select one.)

- Sudden symptoms (fever, pain, injury, etc.)
- A chronic condition (an ongoing illness)
- Mental health problems (low mood, anxiety, trouble sleeping, etc.)
- Test results (health checkups, blood tests, imaging, etc.)
- Medicines (how to take them, side effects, interactions, etc.)
- Other (free text)

**Q9.** Which of the following describes the symptom or condition you asked about? (Select one.)

- A new symptom
- A symptom or condition that had continued from before
- Neither (such as general health management)

**Q10.** At about what time did you ask? (Select one.)

- Morning (6:00-12:00)
- Afternoon (12:00-18:00)
- Evening (18:00-24:00)
- Late night (0:00-6:00)
- Don’t remember

**Q11.** Which best describes how that use related to visiting a medical facility? (Select one.)

- I used it before a visit (for example, I was unsure whether to visit or was preparing for a visit)
- I used it after a visit (for example, I wanted to understand an explanation or test results)
- I was not considering a visit

**Q12.** After using conversational AI, what did you do about the matter? If you used it about someone close to you, include what that person did. (Select all that apply.)

- Visited a medical facility
- Decided not to visit
- Postponed a visit
- Called an ambulance or visited an emergency department
- Called a telephone health advice line (#7119 for emergencies, or #8000 for children)
- Bought or used an over-the-counter medicine
- Changed how I took a prescription medicine, or stopped taking it
- Told a physician or pharmacist what I had found with AI

- Other (free text)
- Did nothing in particular (cannot be selected with other options)

**Q13.** Please write, as far as you remember, what you typed to conversational AI that time. (Free text, up to 300 Japanese characters.)

Text shown below the item: Please do not write anything that could identify a person, such as a name or address.

### Supplementary Material 2. Questionnaires (Japanese original)

Wording as shown on the response screens. The notes after some items (in Japanese) were not shown to respondents.

実際の回答画面で用いた文言を示す。回答はすべて必須で、選択肢の順番は固定した。同意説明文は含めていない。

#### スクリーニング調査（2026 年 10 月 28〜29 日、5 問）

**Q1**　次の中で、あなたが利用したことがあるものをすべてお知らせください。（複数選択）

- チャット GPT：ChatGPT（OpenAI）
- コパイロット：Copilot（Microsoft）
- ジェミニ：Gemini（Google）
- その他生成 AI（サービス名を記入してください）
- 利用したことはない

Q2 の設問文の前に置いた説明：ここからは「対話型 AI」についてうかがいます。対話型 AI とは、ChatGPT、Gemini、Copilot、Claude など、文章で質問すると文章で答える AI のことです。検索結果に表示される AI の要約を読んだだけの場合は含めません。AI の要約から続けて質問した場合（Google の AI モードなど）は含めます。

**Q2**　過去 6 か月の間に、健康についての質問に答えてもらうために、対話型 AI をどのくらい使いましたか。（単一選択）

1. 数回（1〜5 回程度）
2. 月に 1 回
3. 週に 1 回
4. 週に 1 回より多い
5. 対話型 AI を知らない
6. 過去 6 か月は使っていないが、6 か月〜1 年前に使った

7. 1 年以上使っていない（一度も使ったことがない場合も含む）
8. わからない
9. 答えたくない

1～4 を選んだ人を利用者とした。

**Q3**　病院や診療所にかかるとき、問診票などの書類を自分ひとりで記入することに、どのくらい自信がありますか。（単一選択）

- 非常に自信がある
- かなり自信がある
- ある程度自信がある
- 少し自信がある
- まったく自信がない

**Q4**　過去 6 か月の間に対話型 AI に質問したのは、どのようなことについてですか。（複数選択。Q2 で 1～4 を選んだ人のみ）

- 急な症状（熱、痛み、けがなど）
- 持病（続いている病気）
- こころの不調（気分の落ち込み、不安、眠れないなど）
- 検査結果（健康診断、血液検査、画像検査の結果など）
- 薬（飲み方、副作用、飲み合わせなど）
- その他（具体的にお書きください）

**Q5**　過去 6 か月の間に対話型 AI に質問したのは、誰の健康についてですか。（複数選択。Q2 で 1～4 を選んだ人のみ）

- 自分
- 子ども
- 親
- その他の家族
- その他（友人・知人・職場の同僚など具体的にお書きください）

### 本調査（2026 年 11 月 9 日、13 問）

設問の前に置いた文：まず、過去 6 か月の間に、ご自身や身近な人の健康について対話型 AI に質問したことについてうかがいます。

**Q1**　過去 6 か月の間に、次のことのために対話型 AI を使ったことはありますか。（複数選択）

- 自分の症状（または家族や知り合いの症状）が何を意味するのかを知る
- 自分または家族や知り合いの特定の健康問題について、どうすればよいかを知る
- 特定の病気や健康状態について知る
- 健康的な生活習慣について知る
- 自分（または家族や知り合い）の健康を改善する計画を立てる
- 薬、検査、治療についてくわしく知る（安全性、副作用、飲み合わせなど）
- 自分または家族や知り合いが医師にかかるべきかどうかを判断する
- 医学用語の意味を理解する
- 血液検査や画像検査の結果を読み解く
- その他
- わからない（ほかの選択肢と同時には選べない）
- 答えたくない（ほかの選択肢と同時には選べない）

Ayre ら（2025）の設問を翻案した。

**Q2**　対話型 AI が答える内容を、どのくらい信頼していますか。（単一選択）

- 完全に信頼している
- かなり信頼している
- ある程度信頼している
- 少し信頼している
- まったく信頼していない
- わからない
- 答えたくない

Ayre ら（2025）の設問を翻案した。

**Q3**　健康や医療に関する情報について、次のそれぞれが伝える内容を、どのくらい信頼していますか。（項目ごとに単一選択）

項目（この順番で表示）

- 対話型 AI が答える内容
- 医師などの医療職の説明
- テレビ（ニュースや健康番組）が伝える内容
- 新聞（紙・電子版）の記事
- ネットのニュース（Yahoo!ニュースなど）
- インターネット検索で見つけたサイト

- SNS や動画サイト（X、YouTube など）の投稿

選択肢は Q2 と同じ 7 つ。本研究で作成した設問である。

**Q4**　からだの健康（こころの健康に関するものは除く）について対話型 AI を使ったあと、医師などの医療職に相談しましたか。（単一選択）

- はい、相談した
- いいえ、相談しなかった
- からだの健康については使っていない

KFF（2026）の設問を翻案した。

**Q5**　健康に関する情報を得るために対話型 AI を使う理由は、人によってさまざまです。次のそれぞれについて、あなたが健康に関する情報を得るために対話型 AI を使った理由として、主な理由だったか、理由の一つだったか、理由ではなかったかをお答えください。（項目ごとに単一選択）

項目（この順番で表示）

- かかりつけの医師などがいない、または受診の予約が取れなかった
- 受診にかかる費用を払う余裕がなかった
- 受診するかどうか決める前に、調べておきたかった
- 健康に関することを、人に知られずに調べるほうが気が楽だった
- この項目は、必ず「理由の一つだった」を選んでください（注意確認の項目）
- 対話型 AI の情報は、医師などの説明と同じくらい信頼できると感じた
- 医師などと話す前に、検査結果を受け取った
- すぐに情報や支えがほしかった

選択肢：主な理由だった／理由の一つだった／理由ではなかった。KFF（2026）の設問を翻案した（5 番目の項目を除く）。

設問の前に置いた文：ここからは、対話型 AI に健康について質問した、いちばん最近の 1 回についてうかがいます。

**Q6**　そのとき主に使ったサービスはどれですか。（単一選択）

- ChatGPT
- Gemini（アプリまたはサイト）
- Copilot
- Claude

- Google 検索の AI モード（検索結果の AI による概要から続けて質問した場合を含む）
- その他（記入欄あり）

**Q7**　誰の健康についての質問でしたか。（単一選択）

- 自分
- 子ども
- 親
- その他の家族
- その他（友人・知人など）（記入欄あり）

**Q8**　どのようなことについての質問でしたか。（単一選択）

- 急な症状（熱、痛み、けがなど）
- 持病（続いている病気）
- こころの不調（気分の落ち込み、不安、眠れないなど）
- 検査結果（健康診断、血液検査、画像検査の結果など）
- 薬（飲み方、副作用、飲み合わせなど）
- その他（記入欄あり）

**Q9**　質問した症状や病気は、次のどれにあたりますか。（単一選択）

- 新しく出てきた症状
- 以前から続いている症状や病気
- どちらでもない（健康管理についてなど）

**Q10**　質問したのは、何時ごろでしたか。（単一選択）

- 朝（6 時～12 時）
- 昼（12 時～18 時）
- 夜（18 時～24 時）
- 深夜（0 時～6 時）
- 覚えていない

**Q11**　そのときの、医療機関の受診との関係にいちばん近いものはどれですか。（単一選択）

- 受診する前に使った（受診するか迷っていた、受診の準備をしていたなど）
- 受診したあとに使った（説明や検査結果を理解したかったなど）
- 受診は考えていなかった

**Q12**　対話型 AI を使ったあと、その件でしたことをすべてお選びください。身近な人のことで使った場合は、その人がしたことを含みます。（複数選択）

- 医療機関を受診した
- 受診するのをやめた
- 受診を先延ばしにした
- 救急車を呼んだ、または救急外来を受診した
- 救急安心センター（#7119）や小児救急電話相談（#8000）に相談した
- 市販薬を買った、または使った
- 処方された薬の飲み方を変えた、または飲むのをやめた
- 医師や薬剤師に、AI で調べたことを話した
- その他（記入欄あり）
- 特に何もしなかった（ほかの選択肢と同時には選べない）

**Q13**　そのとき対話型 AI に打ち込んだ文章を、覚えている範囲で書いてください。（自由記述、300 字以内）

設問の下に置いた文：氏名・住所など、個人を特定できる情報は書かないでください。
